# A prospective evaluation of the analytical performance of GENECUBE^®^ HQ SARS-CoV-2 and GENECUBE^®^ FLU A/B

**DOI:** 10.1101/2021.02.24.21252337

**Authors:** Yoshihiko Kiyasu, Yusaku Akashi, Akio Sugiyama, Yuto Takeuchi, Shigeyuki Notake, Asami Naito, Koji Nakamura, Hiroichi Ishikawa, Hiromichi Suzuki

## Abstract

**Background:** Molecular tests are the mainstay for detecting severe acute respiratory syndrome coronavirus 2 (SARS-CoV-2). However, their accessibility can be limited by the long examination time and inability to evaluate multiple samples at once. This study evaluated the analytical performance of the newly developed rapid molecular assays GENECUBE^®^ HQ SARS-CoV-2 and the GENECUBE^®^ FLU A/B.

**Method:** This prospective study was conducted between December 14, 2020, and January 9, 2021, at a polymerase chain reaction (PCR) center. Samples were collected from the nasopharynx with flocked swabs. Molecular tests were performed with the GENECUBE^®^ system and reference reverse transcription (RT)-PCR, and the results of the two assays were compared.

**Result:** Among 1065 samples, 81 (7.6%) were positive for SARS-CoV-2 on the reference RT-PCR. Three showed discordance between GENECUBE^®^ HQ SARS-CoV-2 and the reference RT-PCR; the total, positive and negative samples of concordance for the two assays were 99.7%, 100%, and 99.7%, respectively. All discordant cases were positive for GENECUBE^®^ HQ SARS-CoV-2 and negative for the reference RT-PCR. SARS-CoV-2 was detected from all three samples by another molecular assay for SARS-CoV-2. For the GENECUBE^®^ FLU A/B, the total, positive and negative samples of concordance for the two assays were 99.5%, 100%, and 99.1%.

**Conclusion:** The GENECUBE^®^ HQ SARS-CoV-2 and GENECUBE^®^ FLU A/B demonstrated sufficient analytical performance to detect SARS-CoV-2 and influenza virus A/B.

**Key points:** We prospectively evaluated the analytical performance of the newly developed rapid molecular assays GENECUBE^®^ HQ SARS-CoV-2 and the GENECUBE^®^ FLU A/B. The two assays showed >99% concordance rate compared with a reference PCR, which indicated their sufficient analytical performance to detect SARS-CoV-2 and influenza virus A/B.

## 1 Introduction

The severe acute respiratory syndrome coronavirus 2 (SARS-CoV-2), which causes coronavirus disease 2019 (COVID-19), has rapidly spread worldwide [1]. The healthcare system was globally inundated with COVID-19 patients and suffered a detrimental burden [1]. During the pandemic, the timely and accurate identification of patients with SARS-CoV-2 is essential for early isolation and treatment [2].

Several guidelines have recommended nucleic acid amplification tests (NAATs) be performed for SARS-CoV-2 testing due to their high sensitivity and specificity [3,4]. However, it usually takes several hours to days to obtain results for NAATs [5], and few NAATs can concurrently detect other respiratory viruses sharing similar manifestations as SARS-CoV-2 [6]. These drawbacks can limit their accessibility and utility in the clinical setting.

GENECUBE^®^ (TOYOBO Co., Ltd., Osaka, Japan) is a rapid, fully automated genetic analyzer that uses the Qprobe-PCR method [7]. The system automatically prepares reaction mixtures and amplifies and detects target genes in a short time, which, in a single run, can analyze up to eight samples and four items at the same time. GENECUBE^®^ has been applied for several pathogens, including *Mycobacterium tuberculosis* [8], *M. avium, M. intracellulare, Neisseria gonorrhoeae* [9], *Chlamydia trachomatis* [9], *Mycoplasma pneumoniae* [7,10], *Staphylococcus aureus (nuc* and *mecA)* [11] and the toxin gene of *Clostridiodes difficile* [12].

To evaluate the SARS-CoV-2 and influenza A/B virus, the authors of the present study (HS and AS) created two new molecular assays named the GENECUBE^®^ HQ SARS-CoV-2 and GENECUBE^®^ FLU A/B, which were approved in Japan in October and December 2020, respectively. The GENECUBE^®^ HQ SARS-CoV-2 and GENECUBE^®^ FLU A/B can be performed simultaneously or independently within approximately 25 minutes using purified or heat-inactivated respiratory samples.

In this study, we evaluated the analytical performance of GENECUBE^®^ HQ SARS-CoV-2 and the GENECUBE^®^ FLU A/B.

## 2 Methods

A prospective comparison between the GENECUBE^®^ examination and reference real-time polymerase chain reaction (PCR) method was performed with nasopharyngeal samples obtained between December 14, 2020, and January 9, 2021, at a PCR center in Tsukuba, Ibaraki Prefecture, Japan. During the COVID-19 endemic period, sample-collecting for PCR in the Tsukuba district was intensively performed with a drive-through-type method at the PCR center in Tsukuba Medical Center Hospital (TMCH) following a referral from a local public health center and 63 primary care facilities and among healthcare workers at TMCH. Due to the low prevalence rate of influenza and lack of fresh influenza virus-positive samples, we used preserved Universal Transport Medium™ (UTM™) samples that had been stored at −80 °C and used in our previous study [13].

The ethics committee of TMCH approved the present study (approval number: 2020-046), and informed consent was obtained from patients for their participation in the part of the current research using fresh samples.

### 2.1 Sample collection and procedures for comparisons

For sample collections, we obtained a nasopharyngeal sample for PCR with FLOQSwab™ (Copan Italia S.p.A., Brescia, Italy) as previously described [14]. The swab sample was diluted in 3 mL of UTM™ (Copan Italia S.p.A.), and the UTM™ was then transferred to a microbiology laboratory located next to the drive-through sample-collecting place of the PCR center.

After obtaining the UTM™ samples, purification and RNA extraction were performed with a magLEAD 6gC (Precision System Science Co., Ltd., Chiba, Japan) from 200-µL aliquots of UTM™, and the RNA was eluted in 100 µL for GENECUBE^®^ and reference real-time reverse transcription (RT)-PCR using the LightCycler^®^ 96 Real-Time PCR System (Roche, Basel, Switzerland) and a method developed by the National Institute of Infectious Diseases (NIID), Japan for SARS-CoV-2 [15]. The quality of extraction compared with the QIAamp^®^ Viral RNA Mini Kit (QIAGEN N.V., Hilden, Germany) is summarized in Supplementary Table 1. In the present study, a duplicate analysis for N2 genes was performed for the evaluation of SARS-CoV-2.

If discordance was recognized between GENECUBE^®^ and the reference real-time RT-PCR, a re-evaluation was performed with an Xpert^®^ Xpress SARS-CoV-2 and GeneXpert^®^ (Cepheid Inc., Sunnyvale, CA, USA) for SARS-CoV-2 [16] and with the Biofire^®^ FilmArray^®^ Respiratory Panel version 2.1 (BioFire Diagnostics, Salt Lake City, UT, USA) for the influenza virus [17]. As an additional comparison, heat extraction was performed for GENECUBE^®^ HQ SARS-CoV-2 and compared with the reference real-time RT-PCR in this study.

### 2.2 The GENECUBE^®^ assay evaluation with the GENECUBE^®^ HQ SARS-CoV-2 and GENECUBE^®^ FLU A/B

The GENECUBE^®^ can analyze SARS-CoV-2 and the influenza virus simultaneously or independently based on the examiner’s request using the GENECUBE^®^ HQ SARS-CoV-2 and GENECUBE^®^ FLU A/B. The RT-PCR conditions were as follows: reverse transcription reaction 42 °C for 2 min, denaturation at 97 °C for 15 s, and 50 cycles of 97 °C for 1 s, 58 °C for 3 s and 63 °C for 5 s. The RT-PCR products were subjected to a melting point analysis, the conditions of which were as follows: 94 °C for 30 s and 39 °C for 30 s, followed by heating from 40 °C to 75 °C in increments of 0.40 °C/s. Data were analyzed automatically and displayed on the GENECUBE^®^ monitor after completion of the assay evaluation.

For the GENECUBE^®^ examination with the heat extraction method, 100-µL aliquots of UTM™ were mixed with 10 μL of protease K solution (Kanto Chemical Co., Inc., Tokyo, Japan) and heated at 65 °C for 5 minutes and 95 °C for 5 minutes. The inactivated samples were diluted with an equal volume of lysis buffer and used for the assay evaluation.

### 2.3 Evaluation of the limit of detection (LOD) and the feasibility of pooled testing for the GENECUBE^®^ HQ SARS-CoV-2

To evaluate the LOD, twelve fresh SARS-CoV-2-positive UTM™ samples obtained within three days before the evaluation were measured for their viral loads and diluted with UTM™ to approximate concentrations of <100 copies/test and <10 copies/test. In total, 24 positive samples were prepared for the evaluation of LODs and measured in quadruplicate for three examination methods: the reference RT-PCR method, GENECUBE^®^ with the automated purification method, and GENECUBE^®^ with the heat extraction method. Fresh positive influenza virus UTM™ samples were not obtained during the study, and the LOD study was not performed for GENECUE^®^ FLU A/B at present.

The feasibility of pooled testing for the GENECUBE^®^ HQ SARS-CoV-2 was also evaluated. A pool size of 5 was chosen according to the protocol for pooled sample testing for COVID-19 established by the Ministry of Health, Labour and Welfare of Japan. For the pooled analysis, 60 pooled samples were prepared: 20 of 5 pooled negative samples, 20 of 4 pooled negative samples and 1 low viral load samples (30 < Ct value < 35), 20 of 4 pooled negative samples, and 1 moderate to high viral load sample (Ct value < 30). The RNA was extracted with a magLEAD 6gC from 200-µL aliquots of UTM™ was eluted in 50 µL, and then purified samples were evaluated by GENECUBE^®^ HQ SARS-CoV-2 and reference real-time RT-PCR.

### 2.4 Statistical analyses

The positive concordance rate, negative concordance rate, and total concordance rate of the GENECUBE^®^ HQ SARS-CoV-2 and GENECUBE^®^ FLU A/B compared with reference real-time RT-PCR were calculated using the Clopper and Pearson methods with 95% confidence intervals (CIs). All calculations were conducted using the R 3.3.1 software program (The R Foundation, Vienna, Austria).

## 3 Results

### 3.1 Analytical sensitivity of the GENECUBE^®^ HQ SARS-CoV-2

The results of the LOD evaluation are summarized in Table 1. The reference RT-PCR correctly detected up to 5 copies/test, while GENECUBE^®^ with the automated extraction method detected up to 6 copies/test determined by the reference RT-PCR. The LOD of GENECUBE^®^ with the heat extraction method was 17 copies/test determined by the reference RT-PCR.

**Table 1.**
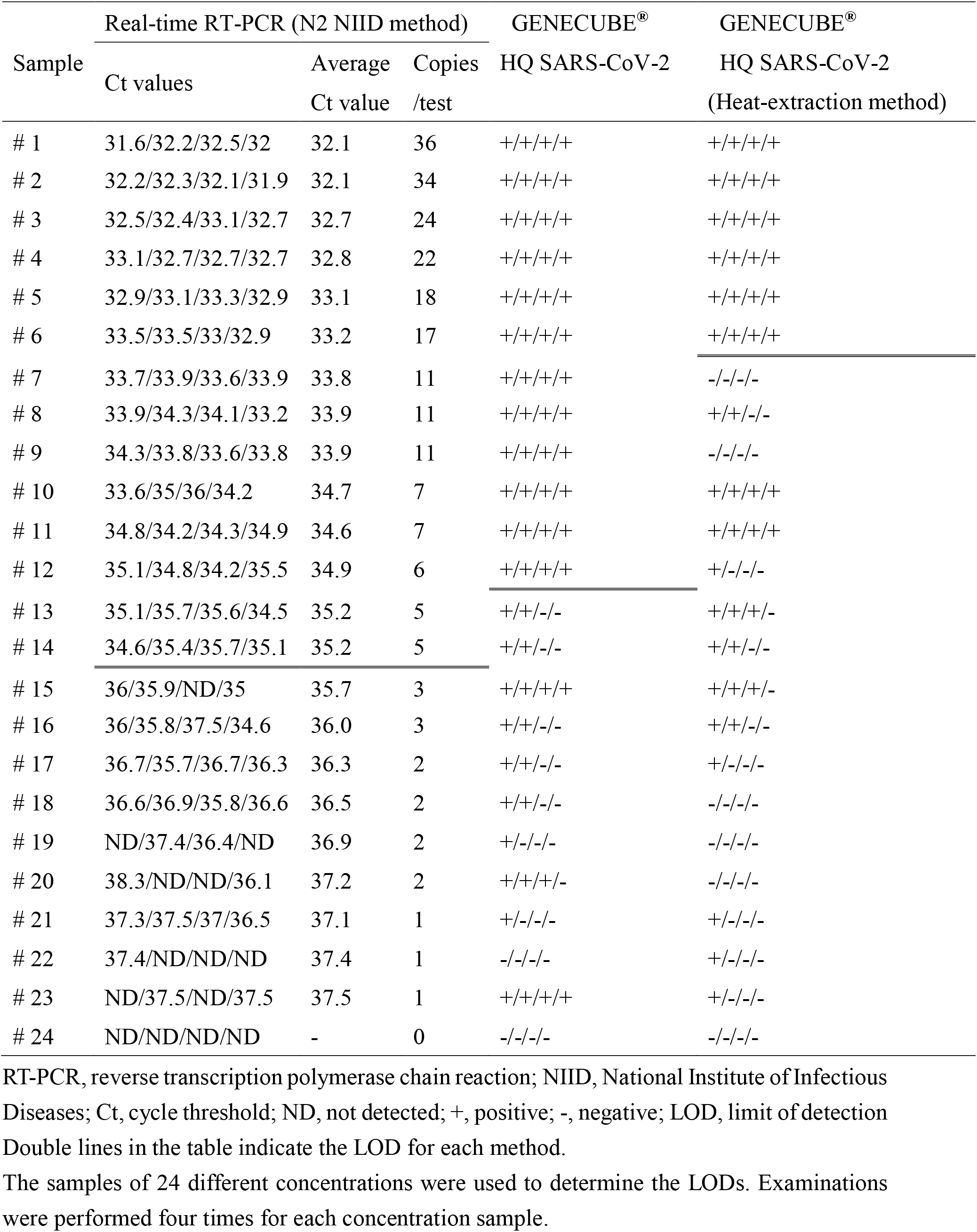
The LOD test results of three SARS-CoV-2 detection methods

### 3.2 A comparison of the GENECUBE^®^ HQ SARS-CoV-2 with reference RT-PCR

During the study period, we evaluated 1065 nasopharyngeal samples. Of these 1065 samples, 486 (45.6%) were obtained from symptomatic patients, and 579 (54.4%) were obtained from asymptomatic patients.

The comparison of the GENECUBE^®^ HQ SARS-CoV-2 assay with the reference RT-PCR for purified samples is summarized in Table 2a. The total, positive and negative concordance of the two assays were 99.7% (1062/1065), 100% (81/81), and 99.7% (981/984), respectively. Of the three samples with discordance between the two assays, all samples were negative by the reference RT-PCR assay and positive by the GENECUBE^®^ assay. SARS-CoV-2 was detected from all three samples by the GeneXpert^®^ for SARS-CoV-2 (Table 3).

**Table 2a.**
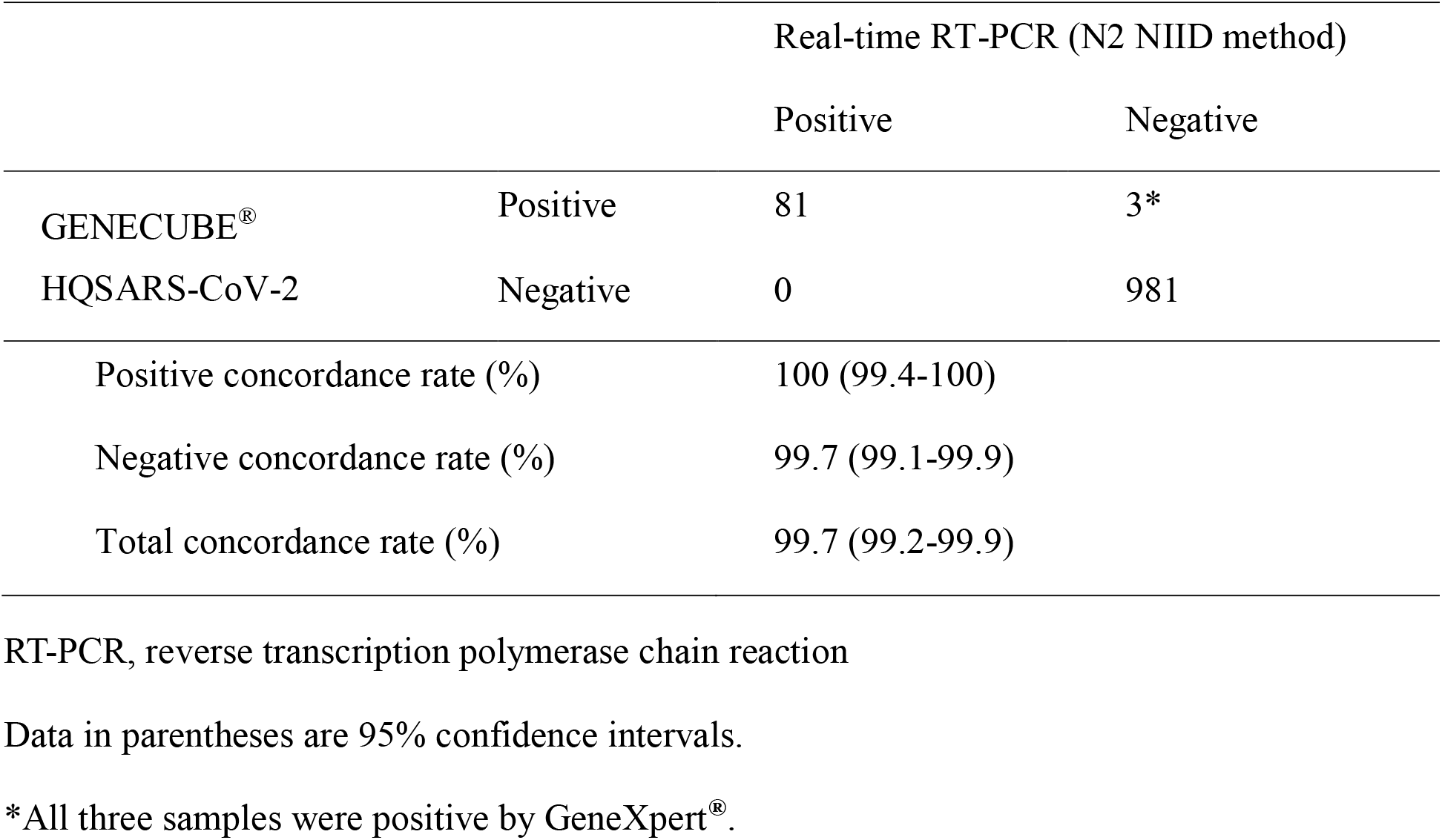
Concordance rate of the GENECUBE^®^ HQ SARS-CoV-2

The comparison of the GENECUBE^®^ HQ SARS-CoV-2 assay with the heat extraction method with the reference RT-PCR is summarized in Table 2b. The total, positive and negative concordance of the two assays were 99.7% (1062/1065), 97.5% (79/81), and 99.9% (983/984), respectively. Of the three samples with discordance between the two assays, one sample was negative by the reference RT-PCR assay and positive by the GENECUBE^®^ assay with the heat extraction method. SARS-CoV-2 was detected from the sample by the GeneXpert^®^for SARS-CoV-2. The other two samples were positive by the reference RT-PCR assay and negative by the GENECUBE^®^ assay with the heat extraction method (Table 3).

**Table 2b.** Concordance rate of the GENECUBE^®^ HQ SARS-CoV-2 with the heat-extraction method

|  |  | Real-time RT-PCR (N2 NIID method) |  |
| --- | --- | --- | --- |
|  |  | Positive | Negative |
| GENECUBE<sup>®</sup> | Positive | 79 | 1 * |
| HQ SARS-CoV-2<br>(Heat-extraction method) | Negative | 2 | 983 |
| Positive concordance rate (%) |  | 97.5 (91.4-99.7) |  |
| Negative concordance rate (%) |  | 99.9 (99.4-100) |  |
| Total concordance rate (%) |  | 99.7 (99.2-99.9) |  |

RT-PCR, reverse transcription polymerase chain reaction
Data in parentheses is 95% confidence intervals.
\*This sample was positive by GeneXpert<sup>®</sup>.

**Table 3.** Detailed data of the five cases with discrepant findings between the three SARS-CoV-2 detection methods

| Case number | Real-time RT-PCR<br>(N2 NIID method) |  | GENECUBE®<br>HQ SARS-CoV-2 | GENECUBE®<br>HQ SARS-CoV-2<br>(Heat-extraction method) | GeneXpert®<br>for SARS-CoV-2 |  |
| --- | --- | --- | --- | --- | --- | --- |
|  | Ct | Ct |  |  | Ct (E) | Ct (N2) |
| 1 | 33.1 | ND | + | - |  |  |
| 2 | ND | ND | + | + | 34.1 | 35.4 |
| 3 | ND | ND | + | - | 34.7 | 37.7 |
| 4 | ND | ND | + | - | 44.7 | 42.7 |
| 5 | 32.9 | 33.4 | + | - |  |  |

RT-PCR, reverse transcription polymerase chain reaction; Ct, cycle threshold; ND, not detected; +, positive; -, negative

### 3.3 Evaluation of the GENECUBE^®^HQ SARS-CoV-2 for pooled samples

The results of the pooling testing with the GENECUBE^®^ HQ SARS-CoV-2 were summarized in Supplementary Table 2a. The GENECUBE^®^ HQ SARS-CoV-2 showed a positive result for every pooled sample with a low viral load (20/20, 100%) and with a moderate-high viral load (20/20, 100%). Meanwhile, the reference RT-PCR assay was negative in 4 low viral load pooled samples.

### 3.4 The comparison of the GENECUBE^®^ FLU A/B with reference RT-PCR

The comparison of the GENECUBE^®^ FLU A/B assay with the reference RT-PCR is summarized in Table 4a. A total of 81 preserved positive UTM™ samples (A: 48 samples, B: 33 samples) were used for the evaluation. Among these 81 positive samples, the sensitivity of antigen testing was 65.4% (53/81).

**Table 4a.**
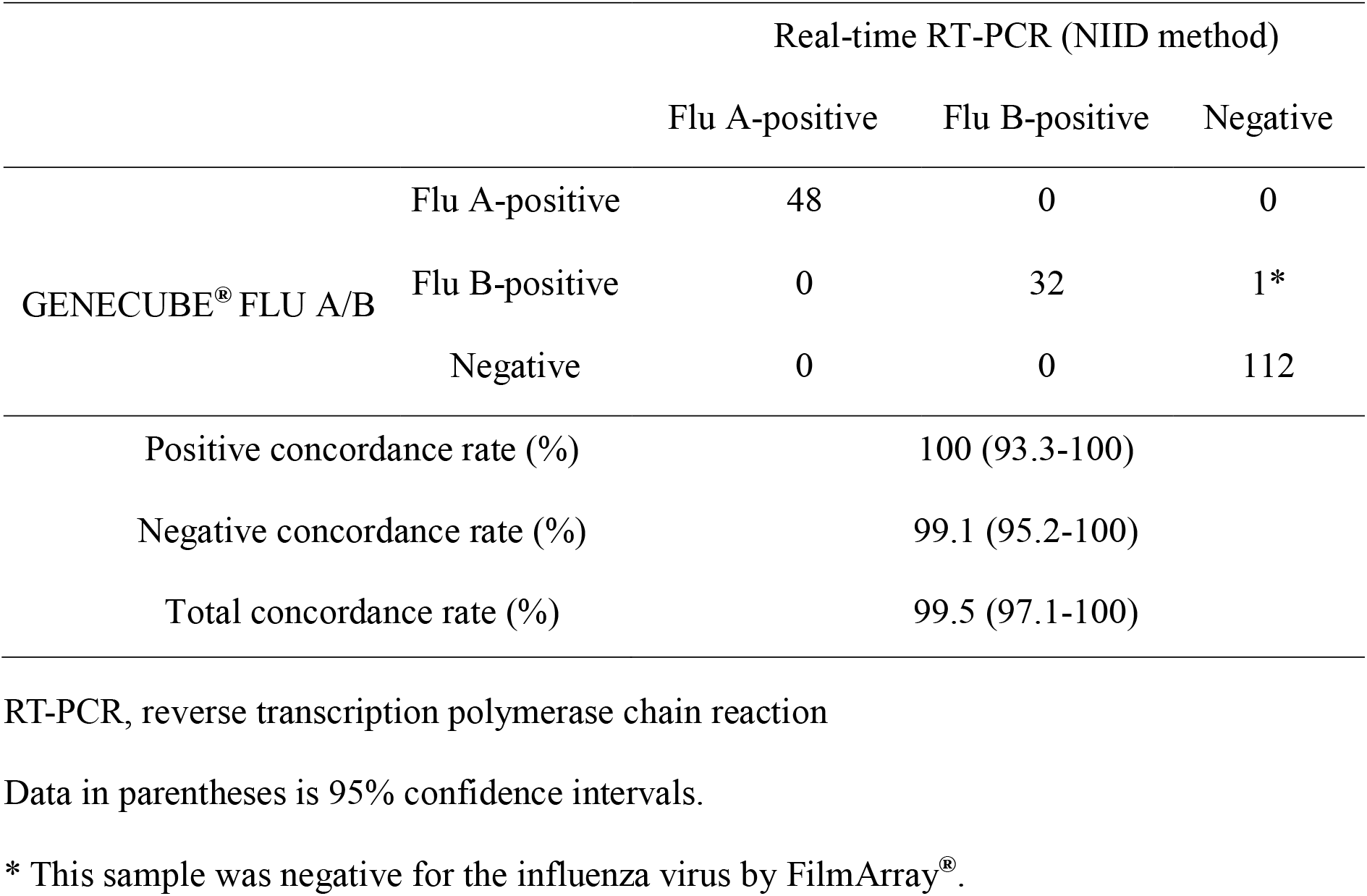
Concordance rate of the GENECUBE^®^ FLU A/B

For purified samples, the total, positive and negative concordance of the 2 assays were 99.5% (191/192), 100% (80/80) and 99.1% (112/113), respectively. One sample with discordance between the two assays was negative by the reference RT-PCR assay and positive by the GENECUBE^®^ FLU A/B assay. The influenza virus was not detected from the sample by FilmArray^®^. (Table 5).

The comparison of the GENECUBE^®^ FLU A/B assay with the heat-extraction method with the reference RT-PCR is summarized in Table 4b. The total, positive and negative concordance of the 2 assays were 97.4% (187/192), 93.8% (75/80) and 100% (113/113), respectively. Of the five samples with discordance between the two assays, all samples were positive by the reference RT-PCR assay and negative by the GENECUBE^®^ FLU A/B assay with heat-extraction method (Table 5).

**Table 4b.** Concordance rate of the GENECUBE^®^ FLU A/B with the heat-extraction method

|  |  | Real-time RT-PCR (NIID method) |  |  |
| --- | --- | --- | --- | --- |
|  |  | Flu A-positive | Flu B-positive | Negative |
| GENECUBE® FLU A/B<br>(Heat-extraction method) | Flu A-positive | 45 | 0 | 0 |
|  | Flu B-positive | 0 | 30 | 0 |
|  | Negative | 3 | 2 | 113 |
| Positive concordance rate (%) |  | 93.8 (86.0-97.9) |  |  |
| Negative concordance rate (%) |  | 100 (95.2-100) |  |  |
| Total concordance rate (%) |  | 97.4 (94.1-99.2) |  |  |

RT-PCR, reverse transcription polymerase chain reaction
Data in parentheses is 95% confidence intervals.

**Table 5.** Detailed data of the six cases with discrepant findings between the three FLU A/B detection methods

| Case<br>number | Real-time RT-PCR<br>(NIID method) |  | GENECUBE®<br>FLU A/B | GENECUBE®<br>FLU A/B<br>(Heat-extraction method) | Notes |
| --- | --- | --- | --- | --- | --- |
|  | Flu A (Ct) | Flu B (Ct) |  |  |  |
| 1 | ND | 31.4 | B | - |  |
| 2 | ND | 34.2 | B | - |  |
| 3 | 24.3 | ND | A | - |  |
| 4 | 31.6 | ND | A | - |  |
| 5 | 29.7 | ND | A | - |  |
| 6 | ND | ND | B | - | Negative for<br>FilmArray® assay |

RT-PCR, reverse transcription polymerase chain reaction; Ct, cycle threshold; ND, not detected; -, negative

## 4 Discussion

In this study, GENECUBE^®^ HQ SARS-CoV-2 and the GENECUBE^®^ FLU A/B showed high analytical performance in this study. The total concordance rate in clinical samples was over 99% between the 2 tests. Regarding the GENECUBE^®^ FLU A/B, the total concordance rate with the reference RT-PCR was 99.5%. The GENECUBE^®^ HQ SARS-CoV-2 provided comparable LODs with the reference RT-PCR and successfully detected SARS-CoV-2 in all positive pooled samples.

The N2 assay of NIID was reported to have a high sensitivity and accuracy for detecting SARS-CoV2 [18]. For the evaluations using purified samples, three discordant cases existed between GENECUBE^®^ HQ SARS-CoV-2 and the reference real-time RT-PCR. Most molecular examinations accurately detect SARS-CoV-2 [21], although some discordance among different systems has been reported [18,22,23]. All our discordant cases tested positive for both GENECUBE^®^ HQ SARS-CoV-2 and GeneXpert^®^; therefore, it appears that these samples indeed contained SARS-CoV-2. When evaluating the pooled samples with a low viral load, we also recognized a similar discordance, GENECUBE^®^ HQ SARS-CoV-2 positive/the reference PCR negative. The sensitivity of molecular examinations may differ between experimental and clinical samples [24]. In this study, we observed the insufficient amplification of the target by the reference real-time RT-PCR (Supplementary Figure 1) and recognized the inhibition which was not indicated in the LODs experiment with diluted UTM™ samples. Multiple factors can influence the results of NAATs, including the quality of the extracted RNA, the presence of RT-PCR inhibitors, genomic mutations [23] and stochasticity observed in very low viral concentrations [17]. These factors may have contributed to the discrepancy in our samples.

Although the extraction methods may influence the RNA recovery [19,20], both rapidity and accuracy are essential for SARS-CoV-2 testing. The heat extraction method can be performed within about 10 minutes, which thus enables the GENECUBE^®^ system to obtain results in less than 40 minutes after the samples arrive at the microbiology laboratory. In this study, GENECUBE^®^ HQ SARS-CoV-2 showed an increase in its LOD when the heat extraction method was used. However, GENECUBE^®^ HQ SARS-CoV-2 with heat extraction showed acceptable analytical performance in clinical specimens, providing a 99.7% concordance rate with the reference real-time RT-PCR.

In the coming winter season, the co-detection of influenza virus will become important for patient management and infection control [25]. The GENECUBE^®^ FLU A/B assay demonstrated only one discordant case with the reference RT-PCR, indicating high analytical performance. The inconsistent case was positive on GENECUBE^®^ FLU A/B but negative on reference RT-PCR and the subsequent Filmarray^®^ assay. This case may be a false positive; however, in our previous study using fresh samples [13], the cobas Liat Influenza A/B assay (Roche Molecular Systems, Pleasanton, CA, USA) provided a positive result for influenza B virus for the discordant sample. Further data are required to evaluate the possibility of false-positive results associated with the GENECUBE^®^ FLU A/B. Similar to GENECUBE^®^ HQ SARS-CoV-2, the sensitivity of GENECUBE^®^ FLU A/B was slightly reduced when using the heat extraction method. Still, the sensitivity of GENECUBE^®^ FLU A/B with heat extraction was comparable to that of other NAATs and far better than that of antigen tests [26].

Several limitations associated with the present study should be mentioned. First, we did not evaluate anterior nasal or saliva samples. Sample collection from the anterior nasal cavity or saliva is less invasive than that from the nasopharynx [27,28], and several molecular examinations can adequately detect SARS-CoV-2 in these samples [28,29]. Further research is warranted to evaluate the performance of GENECUBE^®^ HQ SARS-CoV-2 using samples collected from body sites other than the nasopharynx. Second, the analytical performance may be affected by the future emergence of mutations SARS-CoV-2 and the influenza virus which are involved in the target areas of GENECUBE^®^ HQ SARS-CoV-2 and GENECUBE^®^ FLU A/B. Finally, the data for GENECUBE^®^ FLU A/B were limited due to the low prevalence of the influenza virus, especially for fresh samples.

In conclusion, GENECUBE^®^ HQ SARS-CoV-2 and the GENECUBE^®^ FLU A/B provided high analytical performance and the ability to evaluate multiple samples in a short period of time.

## Data Availability

The data are not publicly available due to their containing information that could compromise the privacy of research participants.

## Acknowledgments

We thank Mrs. Yoko Ueda, Mrs. Mio Matsumoto, and the staff in Tsukuba Medical Center Hospital for their intensive support of this study. Mrs. Yoko Ueda and Mrs. Mio Matsumoto significantly contributed to creating the database of this study.

## Authors’ contributions

Y. Kiyasu drafted the manuscript and performed the statistical analyses. Y. Akashi was the chief investigator and responsible for the manuscript. H. Suzuki supervised the project. A. Sugiyama and A. Naito contributed to the development and execution of molecular assays. Y. Takeuchi, S. Notake, K. Nakamura, and H. Ishikawa supported the preparation of this manuscript. All authors contributed to the writing of the final manuscript.

## Ethics Approval

The present study was performed in line with the principles of the Declaration of Helsinki and with the guidelines of STROBE. The ethics committee of TMCH approved this study (approval number: 2020-046).

