## Supplementary File for "A prospective evaluation of the analytical performance of GENECUBE^®^ HQ SARS-CoV-2 and GENECUBE^®^ FLU A/B"

**Supplementary Table 1a. Comparison of the Ct values between QIAamp® Viral RNA Mini Kit extract and magLEAD extract* for SARS-CoV-2 detection**

| Sample | Real-time RT-PCR (N2 NIID method) | | |
|---|---|---|---|
| | Ct of QIAamp® extract | Ct of magLEAD extract | delta Ct |
| #1 | 26.6 | 26.6 | -0.1 |
| #2 | 24.2 | 25.2 | -1.0 |
| #3 | 20.8 | 21.1 | -0.3 |
| #4 | 19.2 | 18.8 | 0.5 |
| #5 | 13.4 | 13.9 | -0.5 |
| #6 | 16.1 | 16.0 | 0.1 |
| #7 | 12.4 | 15.1 | -2.6 |
| #8 | 16.5 | 17.2 | -0.7 |
| #9 | 18.2 | 18.3 | -0.1 |
| #10 | 22.4 | 22.9 | -0.5 |

The Ct value is the average of the double measurement.

RT-PCR, reverse transcription polymerase chain reaction; NIID, National Institute of Infectious Diseases; Ct, cycle threshold; ND, not detected

*MagDEA® Dx Sv was used for the magLEAD extract.

**Supplementary Table 1b. The comparison of the Ct values between QIAamp® Viral RNA Mini Kit extract and magLEAD extract* for Flu A/B detection**

| Sample | Real-time RT-PCR Flu A or Flu B (NIID method) | | |
|---|---|---|---|
| | Ct of QIAamp® extract | Ct of magLEAD extract | delta Ct |
| #1 (Type A-positive) | 18.5 | 18.9 | -0.4 |
| #2 (Type A-positive) | 21.4 | 21.8 | -0.4 |
| #3 (Type A-positive) | 23.8 | 24.6 | -0.8 |
| #4 (Type B-positive) | 16.3 | 16.3 | -0.1 |
| #5 (Type B-positive) | 22.3 | 21.6 | 0.7 |
| #6 (Type B-positive) | 26.9 | 26.8 | 0.1 |

The Ct value is the average of the double measurement.

RT-PCR, reverse transcription polymerase chain reaction; NIID, National Institute of Infectious Diseases; Ct, cycle threshold; ND, not detected

*MagDEA® Dx Sv was used for the magLEAD extract.

**Supplementary Table 2. The result of pooling method for SARS-Cov-2 detection**

| Sample | Before pooling | | After pooling with 4 negative samples | |
|---|---|---|---|---|
| | GENECUBE® HQ SARS-CoV-2 | Real-time RT-PCR Ct value | GENECUBE® HQ SARS-CoV-2 | Real-time RT-PCR Ct value |
| #1 | + | 12.35 | + | 14.96 |
| #2 | + | 12.38 | + | 13.26 |
| #3 | + | 12.76 | + | 12.47 |
| #4 | + | 14.17 | + | 15.21 |
| #5 | + | 14.46 | + | 14.19 |
| #6 | + | 15.82 | + | 16.65 |
| #7 | + | 16.08 | + | 17.73 |
| #8 | + | 16.33 | + | 17.46 |
| #9 | + | 16.64 | + | 16.72 |
| #10 | + | 16.88 | + | 17.33 |
| #11 | + | 17.69 | + | 19.52 |
| #12 | + | 18.09 | + | 18.72 |
| #13 | + | 18.72 | + | 19.12 |
| #14 | + | 19.22 | + | 19.60 |
| #15 | + | 20.66 | + | 20.62 |
| #16 | + | 20.96 | + | 21.11 |
| #17 | + | 22.38 | + | 23.26 |
| #18 | + | 23.48 | + | 24.87 |
| #19 | + | 23.89 | + | 25.39 |
| #20 | + | 29.40 | + | 30.19 |
| #21 | + | 30.25 | + | 29.92 |
| #22 | + | 30.33 | + | 30.84 |
| #23 | + | 30.67 | + | 29.84 |
| #24 | + | 30.98 | + | 31.69 |
| #25 | + | 31.48 | + | 33.36 |
| #26 | + | 31.54 | + | 32.95 |
| #27 | + | 31.70 | + | 33.13 |
| #28 | + | 31.77 | + | 32.73 |

| | | | | |
|---|---|---|---|---|
| #29 | + | 32.03 | + | 31.05 |
| #30 | + | 32.08 | + | ND |
| #31 | + | 32.25 | + | 32.63 |
| #32 | + | 32.63 | + | 33.32 |
| #33 | + | 32.91 | + | 33.80 |
| #34 | + | 33.22 | + | 30.86 |
| #35 | + | 33.61 | + | 32.96 |
| #36 | + | 33.67 | + | 33.36 |
| #37 | + | 33.74 | + | 34.04 |
| #38 | + | 34.33 | + | ND |
| #39 | + | 34.80 | + | ND |
| #40 | + | 35.00 | + | ND |
| #41 | - | ND | - | ND |
| #42 | - | ND | - | ND |
| #43 | - | ND | - | ND |
| #44 | - | ND | - | ND |
| #45 | - | ND | - | ND |
| #46 | - | ND | - | ND |
| #47 | - | ND | - | ND |
| #48 | - | ND | - | ND |
| #49 | - | ND | - | ND |
| #50 | - | ND | - | ND |
| #51 | - | ND | - | ND |
| #52 | - | ND | - | ND |
| #53 | - | ND | - | ND |
| #54 | - | ND | - | ND |
| #55 | - | ND | - | ND |
| #56 | - | ND | - | ND |
| #57 | - | ND | - | ND |
| #58 | - | ND | - | ND |
| #59 | - | ND | - | ND |
| #60 | - | ND | - | ND |

Ct, cycle threshold; +, positive; -, negative; ND, not detected

**Supplementary Figure 1. The amplification curve of pooled samples on real-time RT-PCR**

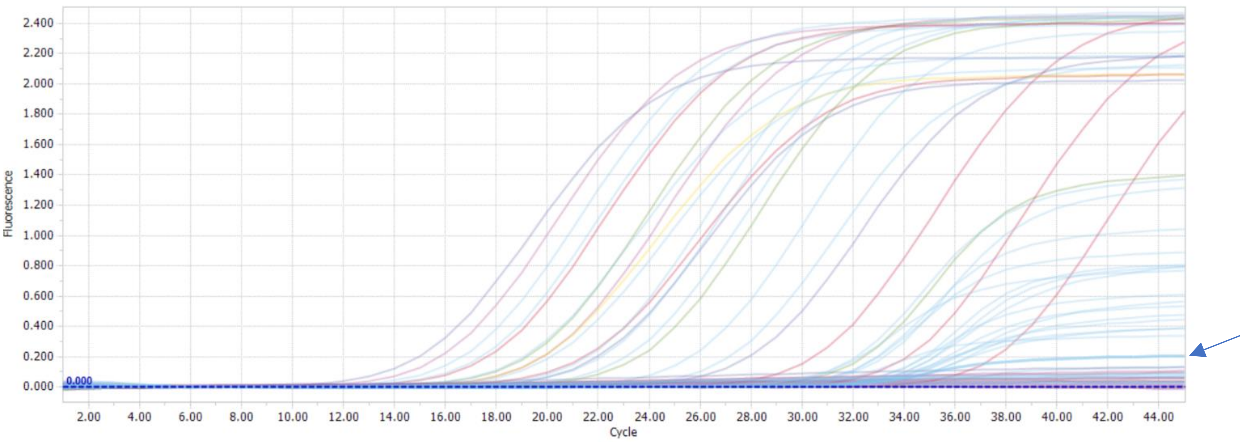


The blue arrow indicates the amplification curve of sample#30 in Supplementary Table 2. The result was judged as negative by LightCycler® 96.
